# Uncertainty-Aware Deep Learning Automates Artifact Correction for Clinical Body Surface Gastric Mapping at Scale

**DOI:** 10.64898/2026.07.08.26357335

**Authors:** Gabriel Schamberg, Nicky Dachs, Hui Yie Teh, Stephen Waite, Chris Varghese, Greg O’Grady, Armen Gharibans

## Abstract

**Objective:** Body surface gastric mapping (BSGM) enables non-invasive measurement of gastric electrophysiology, but signals are approximately 100 times weaker than cardiac potentials and overlap spectrally with motion artifacts, necessitating labor-intensive manual review that limits clinical scalability. We present an uncertainty-aware deep learning framework to automate artifact correction in high-resolution BSGM.

**Methods:** A signal reconstruction network was paired with a parallel uncertainty estimation network, trained on 2,398 multihour, 64- channel recordings from 27 international clinical sites using weak supervision, a physiology-aware loss function, and uncertaintygated quality control.

**Results:** In an independent cohort of 127 patients, per-patient signal reconstruction error was 29% lower than with the industry-standard Wiener filter (10.3% vs 14.6%; lower in 103 patients), and remained lower when both filters retained identical samples (9.8% vs 14.6%). Amplitude– movement correlation was 23% lower. Changing only the filter raised the rhythm index across the cohort and robustly altered automated clinical phenotyping in 7.1% of patients, mostly by removing dysrhythmic classifications from artifact-affected recordings.

**Conclusion:** Uncertainty-aware deep learning enables reliable automated artifact correction in body surface gastric mapping, improving signal fidelity and enabling scalable clinical interpretation.

**Significance:** The system is FDA-cleared (510(k) K252504) and deployed in clinical practice, demonstrating that data-driven artifact correction can meet regulatory requirements for medical devices and reduce dependence on specialist manual review.

## I. Introduction

Functional gastroduodenal disorders affect approximately 10% of the global population [1], yet objective diagnostic tools remain limited [2]. Body surface gastric mapping (BSGM) has recently emerged as a clinically validated, non-invasive platform for measuring gastric myoelectrical activity using high-density electrode arrays [3], [4]. The underlying gastric slow wave dictates the rhythm and propagation of gastric motility at approximately 3 cycles per minute (cpm) [5], and BSGM has demonstrated superior performance in differentiating patient groups and correlating with symptoms compared to legacy methods [6]–[10].

Despite this potential, gastric electrical signals are approximately 100 times weaker than cardiac signals and are highly susceptible to corruption by patient movement, respiration, muscle tension, and electrical activity from adjacent organs. Traditional artifact management is inadequate for BSGM, as simple bandpass filtering cannot separate broadband artifacts from true broadband pathological signals like dysrhythmia, and advanced techniques such as independent component analysis [11] are not well-suited for the complex, variable non-stationarity of BSGM artifacts [12]. Current commercial systems employ automated Wiener filters (WF) [13], but approximately 10% of patients still present with excessive artifacts that the WF cannot adequately resolve, based on internal data. Resolving such cases requires labor-intensive specialist manual review, confining routine clinical use to academic centers [14].

The large volumes of multichannel data generated by BSGM provide a unique opportunity to address this bottleneck with data-driven methods. However, translating deep learning into clinical signal processing requires careful consideration of safety [15], [16]. A particular concern is model hallucination, where a network might confidently generate plausible gastric rhythms from pure noise. Automated systems must therefore provide quantifiable confidence measures [17]. Additionally, models must generalize across the heterogeneity of clinical practice [18], requiring training on large-scale, geographically diverse datasets.

Here we leverage 2,398 high-resolution gastric myoelectrical recordings, each approximately 4 hours in duration, to train models that generalize across diverse clinical populations. We introduce a neural network-based filter (NNF) for automated artifact correction in BSGM that incorporates: (i) an uncertainty-gated dual-branch architecture coupling signal reconstruction with parallel confidence estimation; (ii) a weak supervision training strategy that repurposes an imperfect legacy filter for data labeling, enabling the network to surpass its training source; and (iii) a physiology-aware loss function addressing the extreme dynamic range of gastric electrophysiology. We do not claim that this architecture is optimal. The contribution is a system developed, evaluated and cleared for clinical use at scale. In an independent clinical cohort, these improvements translate to measurable gains in automated phenotyping, recovering gastric rhythms previously obscured by artifact. The system is FDA-cleared (510(k) K252504) and deployed in clinical practice as part of the Gastric Alimetry platform.

## II. Methods

Throughout this paper, we use “training” to describe direct weight updates, “tuning” to describe hyperparameter selection on held-out data, and “testing” to describe evaluation of a locked model on previously unseen data. We avoid the term “validation,” which carries different meanings in machine learning and regulatory contexts.

The end-to-end development and deployment pipeline is shown in Figure 1.

**Fig. 1.**
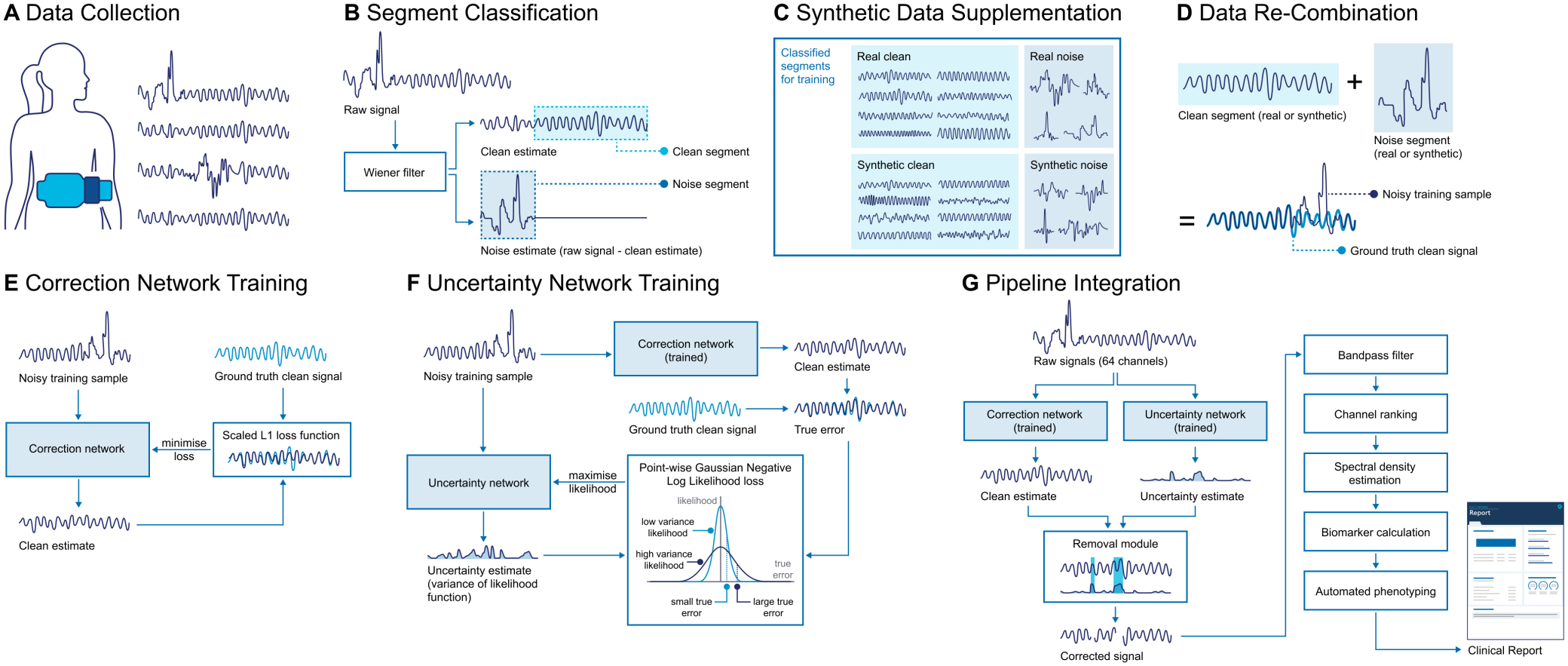
Overview of the data and development pipeline. **A** 64-channel BSGM data is acquired using a high-density electrode array. **B** Recordings are automatically classified into clean and noisy segments. **C** Synthetic clean signals and noise patterns are generated to supplement real training data. **D** Training samples are created by combining clean signals with noise segments drawn from either real or synthetic sources. **E** The correction network is trained to minimize a scaled L1 loss between its output and the ground-truth clean signal. **F** The uncertainty network is trained to maximize the pointwise Gaussian log-likelihood of the true reconstruction error, learning to predict regions where the correction is unreliable. **G** At deployment, raw signals are processed in parallel by both networks; an uncertainty-gated removal module zeros out samples that cannot be reliably reconstructed, and the corrected signal proceeds through bandpass filtering, channel ranking, spectral density estimation, biomarker calculation, and automated phenotyping to generate a clinical report.

### A. Neural Network System Architecture

The system comprises a correction network, for removing artifact components from the BSGM signal, and an uncertainty network that estimates how reliably the correction has been performed. Both networks process each of the 64 channels independently as 1-dimensional convolutional networks that employ variable dilation and skipped connections [19], [20]. The variable dilation enables the network to learn features related both to short-term noise spikes and long-term physiological rhythms while still maintaining modest parameter counts. The architecture is such that the dilations are larger closer to the middle layers of the networks, with skipped connections linking layers with equal dilations at opposing ends of the network. The full network architecture is detailed in Supplementary Fig. 1.

The correction network generates an estimated clean signal in which periods of artifact are replaced by the network’s learned reconstruction of the underlying physiological wave-form. Its final layer uses a linear activation to model the oscillating gastric signal. The parallel uncertainty network outputs a learned estimate of the heteroscedastic aleatoric uncertainty [21], [22] for each time point, quantifying the expected discrepancy between the true signal and the correction network’s output and serving as a per-sample quantitative confidence score. Its final layer uses a Softplus activation, ensuring strictly non-negative outputs appropriate for modeling variance.

The outputs from both networks feed into a final decision-making module. This module normalizes the predicted uncer-tainty by the local signal amplitude (root-mean-square of the corrected signal over a 240-second sliding window), ensuring that small artifacts in low-amplitude signals are treated with equal importance to large artifacts in high-amplitude signals. This normalized uncertainty is compared against a pre-configured threshold *τ* = 1.25 to remove any data that have been unreliably reconstructed (full definition in Supplementary §I). The same threshold is applied at all sites and for all patients. *τ* was selected together with the candidate model on training and tuning data by visual review of spectrograms and per-channel signals, spectral metrics, and the amplitude– movement correlation, prioritizing the elimination of unacceptable outliers in whole recordings while removing about as much data as the WF. The value for *τ* was locked before testing.

### B. Training Paradigm

Each training sample pairs a noisy single-channel input with a known clean target (Section II-E). During preliminary experiments, standard losses (MSE, L1) showed conflicting biases across the dynamic range whereby rare high-voltage outliers (*>* 150 *µ*V, *<*1% of subjects) reconstructed with suppressed amplitude. To resolve this, we introduce an amplitude-normalized scaled-L1 loss that (i) normalizes the error by the ground-truth signal amplitude, so a 1 *µ*V error on a 10 *µ*V signal is weighted like a 10 *µ*V error on a 100 *µ*V signal, and (ii) reduces the penalty at high-noise time points, prioritizing faithful reconstruction of cleaner segments. Crucially, this scaling lives in the loss rather than in preprocessing, avoiding input normalization at inference where the true amplitude may be obscured by noise.

The correction network loss *L*_*c*_ for a single sample comprised of noisy input **x** = [*x*_1_, …, *x*_*T*_], a clean target **y** = [*y*_1_, …, *y*_*T*_], and an estimated clean signal **ŷ** = [*ŷ*_1_, …, *ŷ*_*T*_], is the mean of the element-wise L1 loss across all *T* time steps, scaled at each time by a factor *S*_*t*_ that is inversely related to the local noise magnitude, and normalized by the L2-norm of the true clean output:

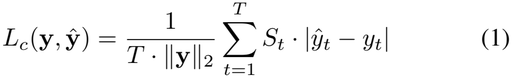

The scaling factor is calculated using the normalized in-stantaneous noise magnitude and two hyperparameters that determine the weighting assigned to the noise magnitude when scaling (*N*_max_) and the maximum allowable reduction (*R*_max_):

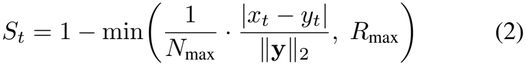

In this study, we set *N*_max_ = 6 and *R*_max_ = 0.9 by design judgment. *N*_max_ sets the normalized noise magnitude at which the reduction would reach 1 if uncapped. With *R*_max_ = 0.9 the scaling saturates at *S*_*t*_ = 0.1 once the normalized noise magnitude reaches *N*_max_*R*_max_ = 5.4. Setting a max reduction of 0.9 ensures that all time points are factored into the loss, with estimation errors at time points for which there is no noise at the input having 10x loss as compared to equivalent errors at time points with large noise magnitude. In practice the weight halves only for artifacts about 60 times the signal RMS (Supplementary Figs. 6 and 7).

The uncertainty network was trained using the Gaussian Negative Log Likelihood (GNLL) loss, which trains the network to predict the variance of the correction network’s error. The loss is minimized when the predicted variance is high where the correction error is large, and low where the error is small. Specifically, letting 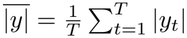 be the uncertainty network output associated with an input **x**, then the uncertainty network loss *L*_*u*_ is given by:

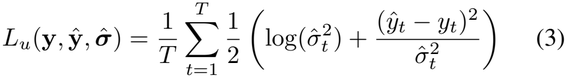

where **y** and **ŷ** are the true clean signal and estimated clean signal (by the correction network) for the same input **x**, respectively.

The networks were trained sequentially. The correction network was trained to convergence, then frozen and used to generate clean-signal estimates that, together with the ground-truth signals, supplied the mean and evaluation point for the uncertainty network’s GNLL loss. Sequential training (rather than jointly optimizing mean and variance estimation with GNLL loss) improved stability and permitted use of the scaled-L1 correction loss. The predicted variance is used only as a thresholded estimate of the local squared error of the frozen correction network, not as a calibrated probabilistic uncertainty, so it affects which data are removed but not the corrected signal.

Two modeling assumptions are involved. First, the recording is modeled as the sum of signal and artifact, as it is for the WF. Second, only the synthetic artifacts, used in half of training samples, are Gaussian. The other half of training samples and all test samples use real artifacts, for which no distribution is assumed.

The models were built and trained using Pytorch [23]. Stochastic gradient descent was performed using the Adam optimizer with a batch size of 32 [24]. 7-minute segments were used for training. For the correction network, the augmented datasets included 1,500,000 segments for training and 30,000 for tuning, trained with a learning rate of 1e-5. The deployed epoch was selected by qualitative review of tuning-set outputs rather than minimum tuning loss. For the uncertainty network, the augmented datasets included 1,000,000 segments for training and 30,000 for tuning, trained with a learning rate of 1e-6.

Both networks were trained on the same mixture of real and synthetic data.

### C. BSGM Data Acquisition System and Protocol

Data were acquired using a non-invasive BSGM system (Gastric Alimetry, Alimetry Ltd., Auckland, New Zealand) during routine clinical use of the Gastric Alimetry system or as part of clinical research studies utilizing the device. All subjects provided informed electronic consent, as outlined in the Alimetry Patient Privacy Notice. Ethical approval for the study was also approved, when required, by institutional review boards in Auckland (New Zealand), Calgary (Canada), and Western Sydney (Australia) (Ethical approvals: AH1130, REB19-1925, and H13541). All procedures involving human subjects were conducted in accordance with the ethical standards of the responsible institutional committees and with the Helsinki Declaration.

The system records from a flexible, single-use patch containing an 8 × 8 grid of 64 electrodes applied to the abdomen over the stomach [3]. The standardized clinical protocol is a continuous recording of at least 4 hours, comprising a 30-minute fasting baseline, a standardized nutrient meal consumed over 10 minutes, and a multihour postprandial recording [25]. Testing was performed only on data collected under this protocol. Tests performed in research studies with custom protocols (e.g. with alternative meals, other simultaneous testing, healthy populations, extended durations, etc.) were also used for model training and tuning so as to ensure exposure to a wide range of signals and artifacts. Extended 24-hour ambulatory recordings, collected under a separate research protocol approved by the Health and Disability Ethics Committee (HDEC; ref: 2024 EXP 21468), were used solely for qualitative demonstration.

### D. Cohorts

The neural networks were developed using 2,398 Gastric Alimetry tests from 27 clinical and research practices across Australia, New Zealand, Canada, the United States, the United Kingdom, Belgium, and Switzerland, partitioned by date into training (*N* = 2,158; 11 March 2020 to 20 November 2023) and tuning (*N* = 240; 22 November 2023 to 6 April 2024) cohorts. The NNF was evaluated on an entirely separate test cohort of 127 patients with chronic gastric symptoms receiving Gastric Alimetry tests as part of their care as prescribed by their gastroenterologist (all tests after 6 April 2024). The test cohort comprised patient data from Australia, New Zealand, Canada, the United States, and the United Kingdom, with 22 of the 127 tests coming from five sites that contributed no training or tuning data. Separation between cohorts was therefore by test and by date. Because patient identifiers are removed during deidentification, patient-level separation could not be verified directly. Repeat testing was estimated at 1.3% of tests, all of it between tests less than 30 days apart, whereas the minimum gap between training and test data was 138 days, so repeat tests are not expected to cross the training–test boundary (Supplementary §V). Detailed demographics, geographical distribution, and baseline recording characteristics are provided in Table I for the testing cohort and Supplementary Table I for the training and tuning cohorts. A subset of patients had BMI *>* 35, which exceeds the Gastric Alimetry system’s instructions for use (IFU) recommendations. These patients were retained in the analysis to reflect the diversity of the clinical population encountered in routine practice. As formal diagnostic classifications were unavailable for this group, we characterized the patients’ clinical presentations using validated measures of gastric symptom severity (Total Symptom Burden Score, scale 0–70 [26]) and psychological comorbidities encompassing stress, depression, and anxiety (Alimetry Gut-Brain Wellbeing Survey, scale 0–40 [27]).

**TABLE 1.** Demographics, Clinical scores, and recording characteristics of the independent test cohort. Continuous variables: Mean*±* SD [Range]; categorical: n (%).

| **Characteristic** | **Test Cohort (n=127)** |
|---|---|
| Age (years) | 35.94 ± 16.14 [14–84] |
| Female sex, n (%) | 101 (80%) |
| BMI (kg/m<sup>2</sup>) | 25.37 ± 7.41 [13.4–54.2] |
| Symptom Burden Score | 21.97 ± 12.81 [5.3–63.2] |
| AGBW Score | 19.10 ± 8.88 [3.0–38.0] |
| United States, n (%) | 39 (31%) |
| New Zealand, n (%) | 35 (28%) |
| Australia, n (%) | 33 (26%) |
| United Kingdom, n (%) | 16 (13%) |
| Canada, n (%) | 4 (3%) |
| Artifact (%) | 25.94 ± 18.31 [0.0–82.7] |
| Impedance (kΩ) | 133.11 ± 94.01 [17.4–468.6] |
| Duration (min) | 240.83 ± 2.68 [240–269] |

### E. Data Sourcing and Preparation

To generate labeled data for training, a version of the existing Gastric Alimetry WF-based artifact detection pipeline, with conservative settings, was applied to each channel to automatically identify and extract segments of “clean” and “noisy” data. Collections of neighboring samples not marked as artifact by the WF were labeled as clean segments and used only if they passed a second-stage WF threshold four times stricter than in clinical use and had electrode impedance below 200 kΩ, a duration of at least 7 minutes, a dominant frequency between 2 and 5 cpm, and at least 75% of power in 2–5 cpm. Collections of neighboring samples marked as artifacts were treated as noise segments, with the noise pattern estimated as the difference between the WF input (noisy data) and output (estimated clean data). This method does not assume the WF is a perfect corrector. Clean segments serve as the ground truth and are therefore selected conservatively, whereas an imperfect noise estimate is sufficient to capture the realistic features of the artifacts the network is designed to correct.

The real-world data were supplemented with synthetically generated data [28]. Synthetic gastric slow waves were modeled as sinusoidal oscillators (1.5–5.0 cpm) with phase drift and amplitude modulation, and synthetic artifacts as transient bursts of 5 seconds to 5 minutes with peak amplitudes reaching orders of magnitude higher than the signal, a subset of which were low-pass filtered to overlap the gastric band. Training samples were constructed by adding a random number of noise segments (0 to 5) to a clean segment, with real and synthetic sources combined equally across four permutations (real/synthetic signal ×real/synthetic noise). Samples with no added noise were included to ensure the network’s ability to preserve clean signals. Supplementing real datasets with synthetic examples can enhance model resilience to out-of-distribution artifacts, even if the synthetic examples are not perfectly realistic [29]. Segment counts and details of the synthetic data are given in Supplementary §IX.

### F. Test Data

Test samples were constructed from the independent test cohort in the same way, using real data only. For each patient recording, segments identified as clean were combined with noise segments from the same patient (0 to 6 per 15-minute sample), with the original clean segment serving as the known “ground truth” against which each filter’s output was measured. This generated a total of 37,177 testing samples. When compared against unprocessed real-world data segments, the testing samples had similar standard deviation, time series complexity, and dominant frequency (Cohen’s *d* = 0.012, −0.036, and −0.071, respectively; Supplementary §II).

### G. Performance Evaluation and Statistical Analysis

The performance of the NNF was directly compared against the existing Gastric Alimetry WF-based artifact detection/correction pipeline, the prior state-of-the-art for commercial BSGM. The primary metric for performance was the Mean Absolute Percentage Error (MAPE), which quantifies the accuracy of the filter’s output signal relative to the ground-truth clean signal. A lower MAPE indicates a more accurate reconstruction. The MAPE was calculated for both the NNF and WF on each segment using only data that had not been removed. Formally, for a true clean signal **y** = [*y*_1_, …, *y*_*T*_] and an estimated signal **ŷ** = [*ŷ*_1_, …, *ŷ*_*T*_], MAPE is defined as:

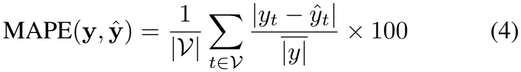

where *V* is the set of time indices where data were not removed by the filter, |*V*| is the number of retained time points, and 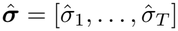 is the mean absolute amplitude of the full true clean signal. This formulation uses a global amplitude denominator rather than pointwise normalization to avoid singularities near zero-crossings inherent in oscillatory physiological signals. The percentage of data removed by each filter was also recorded. Because *τ* was chosen so that the NNF removes about as much data as the WF (Section II-A), this is an operating condition rather than an independent out-come, and MAPE was also compared with removal matched exactly by applying the WF’s artifact mask to both filters. Because samples are clustered within patients, the MAPE and percentage of data removed were compared using per-patient means with Wilcoxon signed-rank tests (*n* = 127), as the paired differences were non-normally distributed, supported by a patient-cluster bootstrap of the sample-level difference. Segment-level means are reported for reference. Generalization was assessed in the 22 tests from unseen sites.

The NNF and WF were also evaluated using the onboard accelerometer on the Gastric Alimetry device. Specifically, the correlation coefficient between the accelerometer’s activity index (root mean square of the variance in the three acceleration axes) and the amplitude of the electrical signal (calculated as a local variance averaged across channels) was calculated for each patient in the independent test cohort. As patient movement often produces high amplitude artifacts, it is assumed that poorer artifact correction will correspond to higher amplitude-movement correlation. A Wilcoxon signed-rank test was used to assess if use of the NNF on the independent testing set reduced the amplitude-movement correlation as compared to the WF. This analysis complements the MAPE and data removal analyses in that it does not rely on classification of segments as clean or noisy.

The performance of the NNF and WF were also visually assessed using both raw signal traces and spectrograms. Raw signal traces serve as a direct side-by-side comparison of the filters, while spectrograms enable evaluation of the effect of the filter differences on the clinical interpretation. Spectrograms were generated using the Gastric Alimetry Algorithm, which comprises a multi-stage signal processing pipeline that includes artifact correction/rejection, filtering, channel selection, and spectral estimation [25]. When comparing spectrograms using the NNF and WF, only the artifact correction/rejection module was modified.

We also evaluated the gating threshold, each filter’s rejection stage, and two untrained comparators (a Hampel filter [30] followed by a 1–6 cpm bandpass, and wavelet shrinkage [31]) (Supplementary §VIII), and stratified test samples into quartiles by the fraction of the true signal’s power in 2–5 cpm to test whether either filter adds rhythm where the true rhythm is weak.

### H. Integration of Filter in Signal Processing Pipeline

The NNF was integrated into the existing Gastric Alimetry signal processing pipeline (Gastric Alimetry Algorithm v3.0.0) as a direct replacement for the previous WF module. The NNF operates on each channel independently. For every input segment, the dual-branch architecture simultaneously generates a reconstructed signal (from the correction network) and a confidence score (from the uncertainty network). The uncertainty score is normalized by the local signal amplitude and the segment is classified as indeterminate and removed from downstream analysis if this normalized value exceeds the pre-configured threshold *τ*. Conversely, if the confidence is within the acceptable range, the raw signal is replaced by the correction network’s output. Following artifact correction, signals undergo bandpass filtering (1–6 cpm) to isolate gastric frequencies, followed by a channel ranking algorithm that identifies the electrodes with the highest signal-to-noise ratio for aggregate analysis.

### I. Spectral Analysis

To generate the clinical spectrograms used for physician interpretation, spectral density estimation was performed on the cleaned data from the top 8 ranked channels. A short-time Fourier transform (STFT) was applied to each channel using a sliding window approach with a window length of 1 minute and a 75% overlap. The resulting time-frequency representations were averaged across the selected channels to produce a single, global patient spectrogram. This aggregation method mitigates the impact of local electrode noise and ensures the visualization reflects the global gastric rhythm.

### J. Automated Phenotyping Evaluation

To evaluate the clinical impact of the NNF, we compared automated phenotyping outcomes between the WF and NNF pipelines for all 127 patients using the Auckland Classification v1.0 (ACv1) [9], running the released algorithm (v3.0.0) twice and changing only the noise filter. ACv1 may assign multiple phenotypes to a single test (Dysrhythmic, High Frequency, Low Meal Response, Sensorimotor, Continuous, Delayed On-set Symptoms, and descriptors such as high- or low-amplitude activity), one or more of which are designated dominant, or may detect none. This classification utilizes standardized spectral biomarkers [32]–[34]: the Principal Gastric Frequency (PGF), the frequency of the most stable oscillation within the gastric range (normative interval 2.65–3.35 cpm); BMI-adjusted amplitude, the average signal amplitude corrected for body mass index (normative interval 22–70 *µ*V); the Gastric Alimetry Rhythm Index (GA-RI), a spectral stability metric (range 0–1; normative threshold ≥0.25); the Meal Response Ratio (MRR), the ratio of average amplitude in the first 2 hours postprandially to the following 2 hours (threshold ≥ 1.0); and symptom–amplitude correlation scores (threshold *>* 0.5). These thresholds are those of the published normative reference intervals, derived from healthy volunteers [33], and of ACv1 [9].

To separate changes that cross a threshold from changes within the variability near it, we defined gray areas around primary thresholds: ±0.02 for GA-RI (0.22 and 0.25), ±0.06 for MRR (1.0), and ±0.06 for symptom–amplitude correlation (0.5). A reclassification was any change in the set of dominant phenotypes. It was deemed robust if the threshold was crossed and at least one paired value was outside an interval around the threshold, and borderline otherwise. Reclassified tests were reviewed visually by the authors, unblinded to filter. Agreement of each biomarker between filters across all 127 tests was summarized by the mean difference with 95% limits of agreement [35] and Lin’s concordance correlation coefficient (CCC) [36].

## III. Results

### A. Quantitative Signal Reconstruction Performance

At the patient level, mean absolute percentage error (MAPE) was lower with the NNF than with the WF in 103 of 127 patients (10.32% vs 14.58%, a 29% relative reduction; *p <* 0.0001; 95% patient-cluster bootstrap interval for the sample-level difference −7.9 to −4.8 percentage points; Table II). Per-patient median MAPE differed less (8.26% vs 8.91%), indicating that the gain is concentrated in the most corrupted samples. The two filters removed a similar amount of data (9.88% vs 11.43%), as intended by the choice of *τ*, and the lower error did not depend on which data were removed. With the WF’s artifact mask applied to both filters, so that identical samples were retained, MAPE was 9.8% with the NNF and 14.6% with the WF (Table III). In the 22 tests from unseen sites, MAPE fell from 13.1% to 10.5% (*p* = 0.023), with no detectable difference in improvement from seen sites (*p* = 0.37). The signals produced by the NNF also had a 23% relative reduction in correlation between gastric amplitude and patient movement as measured by the device’s onboard accelerometer. Notably, this amplitude–movement correlation metric is derived entirely from the onboard accelerometer and is independent of the segment-level labels used for training, providing external validation that the NNF has learned genuine physiological signal characteristics rather than simply replicating the training heuristic.

**TABLE 2.** Results as mean (SD). Patient level: means of per-patient values, Wilcoxon signed-rank (*n* = 127). Sample level (*n* = 37,177 samples) is given for reference.

| **Metric** | **NNF** | **WF** | **p** |
|---|---|---|---|
| *Patient level* |  |  |  |
| MAPE (%) | 10.32 (4.93) | 14.58 (6.03) | <0.0001 |
| Data removed (%) | 9.88 (4.49) | 11.43 (2.12) | <0.0001 |
| Amp.–movement correlation | 0.096 (0.079) | 0.124 (0.085) | <0.0001 |
| *Sample level* |  |  |  |
| MAPE (%) | 9.97 (30.81) | 16.36 (38.46) |  |
| Data removed (%) | 10.22 (11.21) | 11.28 (7.28) |  |

**TABLE 3.** MAPE (%) with and without each filter’s rejection stage, as the mean of per-patient means (*n* = 127). Sample-level values and untrained comparators are given in Supplementary

| **Rejection stage** | **Removed (%)** | **NNF** | **WF** |
|---|---|---|---|
| None | 0 | 16.9 | 27.1 |
| WF artifact mask for both | 11.4 | 9.8 | 14.6 |
| Each filter's own (deployed) | 9.9 / 11.4 | 10.3 | 14.6 |

### B. Rejection Stage, Threshold and Comparators

Table III separates each filter’s correction from its rejection stage. Without a rejection stage the correction network still outperformed the WF (16.9% vs 27.1%; WF better in 6 of 127 patients, *p <* 0.0001), and with removal matched the NNF’s error was about one third lower than the WF’s. A rejection stage therefore matters for both filters, but the NNF’s lower error does not depend on its own gate. The gating threshold traded error against data removed smoothly, from MAPE 2.0% with 86.9% removed (*τ* = 0.25) to 16.9% with none removed (no gating), with no obvious point in the curve indicating optimality (Supplementary Fig. 8). The untrained comparators performed far worse than both filters, with the NNF better in all 127 patients (Supplementary Table III).

### C. Visual Assessment

Fig. 2 provides representative examples of the system’s performance on heavily contaminated signals. The power spectral densities illustrate how the underlying gastric slow wave (∼ 3 cpm) [25] can be overpowered by broadband artifacts. The NNF demonstrates improved performance in both correcting artifacts while preserving the physiological waveform (panel A) and reliably removing irreparably corrupted data (panel B).

**Fig. 2.**
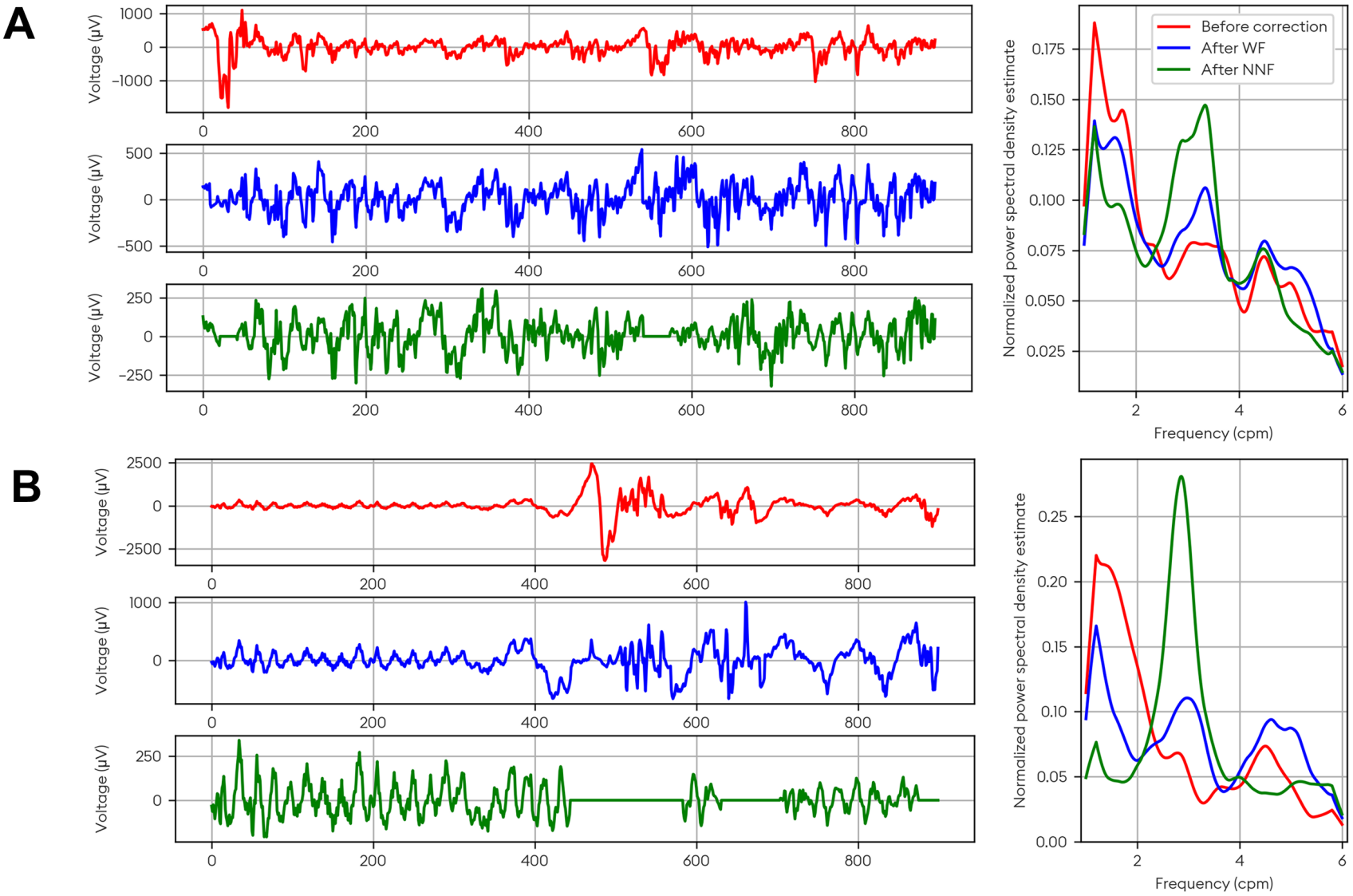
Two 15-minute segments of single channel BSGM recordings. Each panel shows the signal before correction (top), after correction with WF (middle), and after correction with NNF (bottom), with normalized power spectral density estimates (right). Panel A demonstrates improved artifact correction; Panel B demonstrates improved removal of irreparably corrupted data. Flat lines indicate data removed by the filter.

The improvement in signal fidelity has a direct impact on spectral analysis, the central component of BSGM interpretation. In a research protocol where patients stood every 15 minutes for a gastric emptying breath test, movement produced broadband vertical artifacts that obscured the ∼3 cpm rhythm and were only partially removed by the WF; the NNF recovered a clear, stable gastric frequency band (Supplementary Fig. 2). This clean spectral representation is essential for accurate biomarkers such as the Principal Gastric Frequency (PGF) and the Gastric Alimetry Rhythm Index (GA-RI) [33]. The standard Gastric Alimetry test protocol dictates that patients should try to minimize movement. As part of an ongoing research program, data have been collected from patients over a 24-hour period using an ambulatory protocol. In such settings, accurate mechanisms for artifact correction are imperative. Supplementary Fig. 3 demonstrates that the NNF can enable more reliable detection of gastric activity even in settings where patient activity is not monitored or controlled. As compared to the WF, the NNF removes less data, and where data is retained, activity is more concentrated in the gastric activity band.

### D. Impact on Automated Patient Phenotyping

To quantify the translational impact of the NNF on clinical decision-making, we evaluated the impact of improved filtering on automated phenotyping results in the testing cohort (*n* = 127). We applied the algorithmic implementation of the ACv1 [9], the consensus standard for phenotyping patients based on BSGM tests, to both the WF-processed and NNF-processed data.

Changing only the noise filter raised GA-RI by 0.035 on average across the 127 tests, and the other biomarkers remained in close agreement between filters (Table IV). To confirm that this rise does not reflect rhythm added by the network, we analyzed the quartile of test samples with the weakest true 2–5 cpm rhythm. In these samples, the NNF output carried 0.98 of the true gastric-band amplitude, whereas the WF inflated it by 13%. Tests with GA-RI below 0.25 fell from 30 to 20, PGF became detected in 10 tests and was lost in none, and Dysrhythmic labels fell from 25 to 17, with other label counts essentially unchanged.

**TABLE 4.**
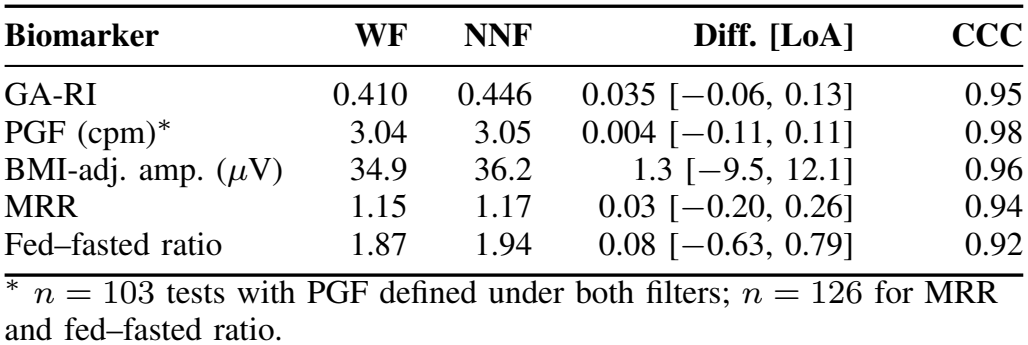
Agreement of overall biomarkers between filters across the 127 test recordings (NNF*−* WF). LoA: 95% limits of agreement; CCC: Lin’s concordance correlation coefficient.

| **Biomarker** | **WF** | **NNF** | **Diff. [LoA]** | **CCC** |
|---|---|---|---|---|
| GA-RI | 0.410 | 0.446 | 0.035 [−0.06, 0.13] | 0.95 |
| PGF (cpm)<sup>*</sup> | 3.04 | 3.05 | 0.004 [−0.11, 0.11] | 0.98 |
| BMI-adj. amp. (*μ*V) | 34.9 | 36.2 | 1.3 [−9.5, 12.1] | 0.96 |
| MRR | 1.15 | 1.17 | 0.03 [−0.20, 0.26] | 0.94 |
| Fed–fasted ratio | 1.87 | 1.94 | 0.08 [−0.63, 0.79] | 0.92 |

<sup>\*</sup> *n* = 103 tests with PGF defined under both filters; *n* = 126 for MRR and fed–fasted ratio.

In total, 20 patients exhibited a change in their dominant automated phenotypes between the WF and NNF pipelines (Supplementary Table II). Of these, 11 were classified as borderline, with changes occurring within the defined bounds. However, 9 patients (7.1% of the independent cohort) demonstrated robust phenotype reclassification.

Across the 9 robustly reclassified tests, GA-RI was 0.201 ± 0.040 with the WF and 0.286 ± 0.027 with the NNF, a mean shift of +0.085 (range: +0.010 to +0.170; Supplementary Fig. 5). These tests were identified in part by GA-RI crossing its threshold, so this shift is descriptive of the subgroup, and the evidence that the NNF raises GA-RI is the difference across all 127 tests (Table IV). In 7 of the 9 cases, the GA-RI shifted from below to above the 0.25 normative threshold. In 8 of the 9 tests, the WF phenotype included a Dysrhythmic classification that was removed or deprioritized by the NNF, while one test was reclassified from Normal (no phenotype detected, with normal spectral metrics and absence of noteworthy symptom profiles) to Sensorimotor after the NNF revealed a previously masked symptom–amplitude association (heartburn correlation: WF = 0.31 vs NNF = 0.52).

The 9 reclassified tests had significantly higher baseline artifact detection rates under the WF (27.1± 9.3%) compared with the remaining 118 tests (15.2± 13.9%; Mann– Whitney *U, p* = 0.001), consistent with these recordings being disproportionately affected by motion-related contamination. Furthermore, the NNF reduced the amplitude–movement correlation by a mean of 0.055± 0.053 across the reclassified tests (WF: 0.178 ±0.073 vs NNF: 0.123 ±0.072; paired *t*-test, *p* = 0.015). This average decrease is roughly double that of the entire testing cohort (0.028, Table II), suggesting that the phenotypic shifts were driven in part by an increased effect of the decoupling of signal from movement artifacts.

Unblinded visual review of the WF and NNF spectrograms agreed with all 9 robust reclassifications (Supplementary Fig. 4). In the clearest cases, the WF-processed spectrograms exhibited pronounced broadband vertical artifacts spanning extended periods of the test, corresponding directly to elevated patient movement. The NNF successfully suppressed these interference patterns to reveal coherent gastric slow wave activity at a physiologically plausible frequency, recovering a stable horizontal band at approximately 3 cpm that was previously obscured. In other cases, the degree of spectral improvement was more subtle but nonetheless sufficient to alter the phenotype classification, with the NNF selectively restoring spectral continuity during contaminated intervals to yield modest but classification-altering improvements in the computed biomarkers.

Across all 9 tests, the directionality of the reclassifications was consistent with the expected effect of artifact removal: un-corrected broadband noise artificially reduces rhythm stability measures (GA-RI), inflates the apparent data loss (reducing the Meal Response Ratio and symptom–amplitude correlations), and can mask the true gastric frequency, leading to false Dysrhythmic phenotypes. The alignment between the recovery of clearly visible rhythmic activity, the reduction in amplitude–movement correlation, and the corresponding shift in classification-driving biomarkers is consistent with a more accurate representation of genuine underlying gastric physiology. No diagnostic reference standard exists for these patients, however, so the correctness of individual reclassifications cannot be evaluated in a clinical context.

## IV. Discussion

This work details the development and validation of a dual neural network system for the automated processing of artifacts in high-resolution BSGM data. The results demonstrate that this data-driven approach is not only significantly more accurate at reconstructing physiological signals but also successfully lowers the barrier to clinical interpretation. By automating the complex quality control tasks previously reserved for highly trained specialists, this technology overcomes a long-standing barrier to the widespread adoption of non-invasive gastric monitoring.

The transition from conventional, parameter-based filters to a learned system represents a fundamental shift in electro-physiological signal processing. Fixed-rule algorithms, such as bandpass or Wiener filters, rely on *a priori* assumptions that are frequently violated by the complex, non-stationary artifacts associated with body surface gastric recordings. The NNF overcomes this by learning the subtle morphological differences between true gastric slow waves and artifacts that mimic them, a capability developed through exposure to thousands of real-world examples. This aligns with recent successes in other medical signal processing domains including computed tomography, electrocardiograms, EEG, and histology [37], [38].

A critical technical innovation of this work is the weak supervision training strategy. We demonstrated that an imper-fect existing filter could be used to generate training data for a neural network that would ultimately outperform the filter used to generate the data. By simplifying the Wiener filter’s task from *correction* to *identification* (i.e., isolating clean vs. noisy segments), we created a large dataset of real-world signals that allowed the NNF to learn signal reconstruction via self-training principles [39]. This was validated by the NNF’s improved ability to decouple gastric amplitude from patient movement. As this metric is completely independent of the training labels, it indicates that the model learned true physiological distinctions rather than simply mimicking the training heuristic.

The system incorporates a dedicated uncertainty quantification module. By separating signal correction from confidence assessment, the system avoids presenting plausible-looking but unreliable reconstructions. The use of Gaussian negative log likelihood (GNLL) loss allows the model to explicitly flag and remove data it cannot reliably reconstruct.

In 9 patients (7.1%), the NNF recovered stable gastric activity from recordings previously compromised by artifact. Because uncorrected broadband noise dilutes the gastric frequency band and artificially lowers the GA-RI [40], the NNF’s separation of noise from signal can prevent false Dysrhythmic assignment. By salvaging these tests, the system reduces reliance on expert review and the rate of non-diagnostic results that delay care.

This study has limitations. First, evaluation was conducted on a single proprietary hardware platform. While the principles are generalizable, performance on different electrode configurations would require separate validation. Second, the “ground truth” for training relied on recombining real signal segments, as simultaneous noise-free recording is physically impossible in this setting. However, we showed that the morphology of recombined signals was similar to that of unmodified signals. Third, the uncertainty is aleatoric only, so inputs unlike the training data are not specifically detected, and noise confined to the gastric band cannot be separated from signal by a single-channel filter. Fourth, the clean training pool was checked only by informal, non-comprehensive visual review. Fifth, no ablation of the loss or architecture was conducted, sensitivity to *N*_max_ and *R*_max_ was not studied, and learned denoisers from other biosignal domains were not compared. Finally, phenotyping lacks a diagnostic reference standard and the visual review was unblinded.

Future work includes epistemic uncertainty and calibrated or patient-adaptive thresholds [41], comparison with learned denoisers, blinded external review of reclassifications, and extension to ambulatory 24-hour monitoring and other body-surface electrophysiology. Potential use cases include BSGM in community and remote settings without specialist review, pediatrics, and longitudinal monitoring.

## V. Conclusion

This work demonstrates that uncertainty-aware deep learning enables reliable automated correction of artifacts in body surface gastric mapping. The dual-network architecture, combining signal reconstruction with parallel uncertainty estimation, achieved a 29% reduction in per-patient reconstruction error and a 23% reduction in amplitude–movement correlation compared with the industry-standard Wiener filter in an independent cohort of 127 patients, and its error remained lower when both filters retained identical data. These signal-level improvements carried through to automated phenotyping, with the system recovering gastric rhythms obscured by artifact and producing robust phenotype reclassifications in 7.1% of patients.

By improving signal fidelity and enabling scalable interpretation of gastric electrophysiology, this framework supports the broader clinical adoption of non-invasive gastric monitoring. The system removes the requirement for specialist manual review at each clinical site, lowering the barrier to entry for generalist clinicians and reducing the rate of non-diagnostic results. The system is FDA-cleared (510(k) K252504) and currently deployed in clinical practice, demonstrating a complete translational pathway from algorithm development through regulatory approval. The weak supervision training strategy and physiology-aware loss function presented here may be applicable to artifact correction challenges in other surface electrophysiology domains where ground-truth clean signals are unavailable.

## Data Availability

The clinical data used in this study were collected under informed consent that permits research use but not unrestricted public release. De-identified summary statistics and aggregate results supporting the findings of this study are provided in manuscript. Individual-level de-identified data may be made available from the corresponding author upon reasonable request, subject to appropriate data use agreements.

## Author Contributions

G.S. and A.G. conceived the study. G.S. and H.Y.T. designed and trained the neural network models. G.S. and N.D. performed the analysis and wrote the manuscript. G.S. and S.W. contributed to data curation, processing pipeline development, and quality assurance. C.V. and G.O.G. developed the Auckland Classification v1.0 and contributed to clinical interpretation. G.O.G. and A.G. supervised the study and secured funding. All authors reviewed and approved the final manuscript.

