## Supplementary Information for "Uncertainty-Aware Deep Learning Automates Artifact Correction for Clinical Body Surface Gastric Mapping at Scale"

#### I. Network Architecture

Both the correction and uncertainty networks are 1-dimensional convolutional networks with identical U-shaped architectures (9 layers each, kernel size $k$=25, same padding) but distinct output activations. Both networks independently process single-channel BSGM input ($\mathbf{x} \in \mathbb{R}^T$). The downstream path uses increasing dilation rates ($d$), expanding the receptive field to capture both short-duration artifact spikes and long-period gastric rhythms. In the upstream path, each convolutional layer's output is summed element-wise with the corresponding encoder layer's pre-activation output (dashed skip connections), preserving fine-grained temporal features. The correction network outputs an estimated clean signal $\hat{\mathbf{y}}$ via a linear activation, while the uncertainty network outputs pointwise aleatoric uncertainty $\hat{\boldsymbol{\sigma}}$ via a Softplus activation. An uncertainty gating module compares each sample's predicted standard deviation to the local signal power, computed as the root-mean-square of the corrected signal over a sliding window of width $w$; samples for which $\hat{\sigma}_t/\text{RMS}_w(\hat{\mathbf{y}})_t > \tau$ are rejected from downstream analysis. The full architecture is shown in Fig. 1.

The gating module operates per channel with a centered window of $w = 240$ s and a threshold of $\tau = 1.25$, used for every channel, patient and site. Both values were selected with the candidate model on training and tuning data, by visual review of whole-recording spectrograms and per-channel signals, spectral metrics and the amplitude–movement correlation. The primary goal was to eliminate unacceptable outliers while removing about as much data as the WF. Both values were locked before testing. Their effect on error and data removed is characterized in Section VIII.

#### II. Augmented Data Validation

To verify that the augmented test samples (generated by recombining clean and noisy segments) were representative of real-world recordings, three signal characteristics were compared between the augmented samples and a reference set of unprocessed single-channel segments drawn from the same patients without regard to clean/noisy classification. For each characteristic, Cohen's $d$ was calculated as $d = (\bar{x}_1 - \bar{x}_2)/s_p$, where $\bar{x}_1$ and $\bar{x}_2$ are the group means and $s_p = \sqrt{(s_1^2 + s_2^2)/2}$ is the pooled standard deviation.

The first characteristic was the standard deviation of each segment, capturing overall signal amplitude variability. The second was time series complexity, quantified using the complexity-invariant distance estimate (CID_CE) [1]. For a segment $\mathbf{s} = [s_1, \dots, s_N]$, the signal is first standardized to zero mean and unit variance, yielding $\tilde{\mathbf{s}}$, and the complexity is then defined as:

$$\text{TSC}(\mathbf{s}) = \sqrt{\sum_{n=1}^{N-1} (\tilde{s}_{n+1} - \tilde{s}_n)^2} \quad (1)$$

This metric increases with the number and magnitude of directional changes in the signal, providing a measure of waveform irregularity independent of amplitude and offset. The third characteristic was the dominant frequency, defined as the frequency corresponding to the peak of the power spectral density estimated via Welch's method [2] with a segment length of 4 minutes. The testing samples had similar standard deviation, time series complexity, and dominant frequency to unprocessed real-world segments (Cohen's $d$ = 0.012, −0.036, and −0.071, respectively).

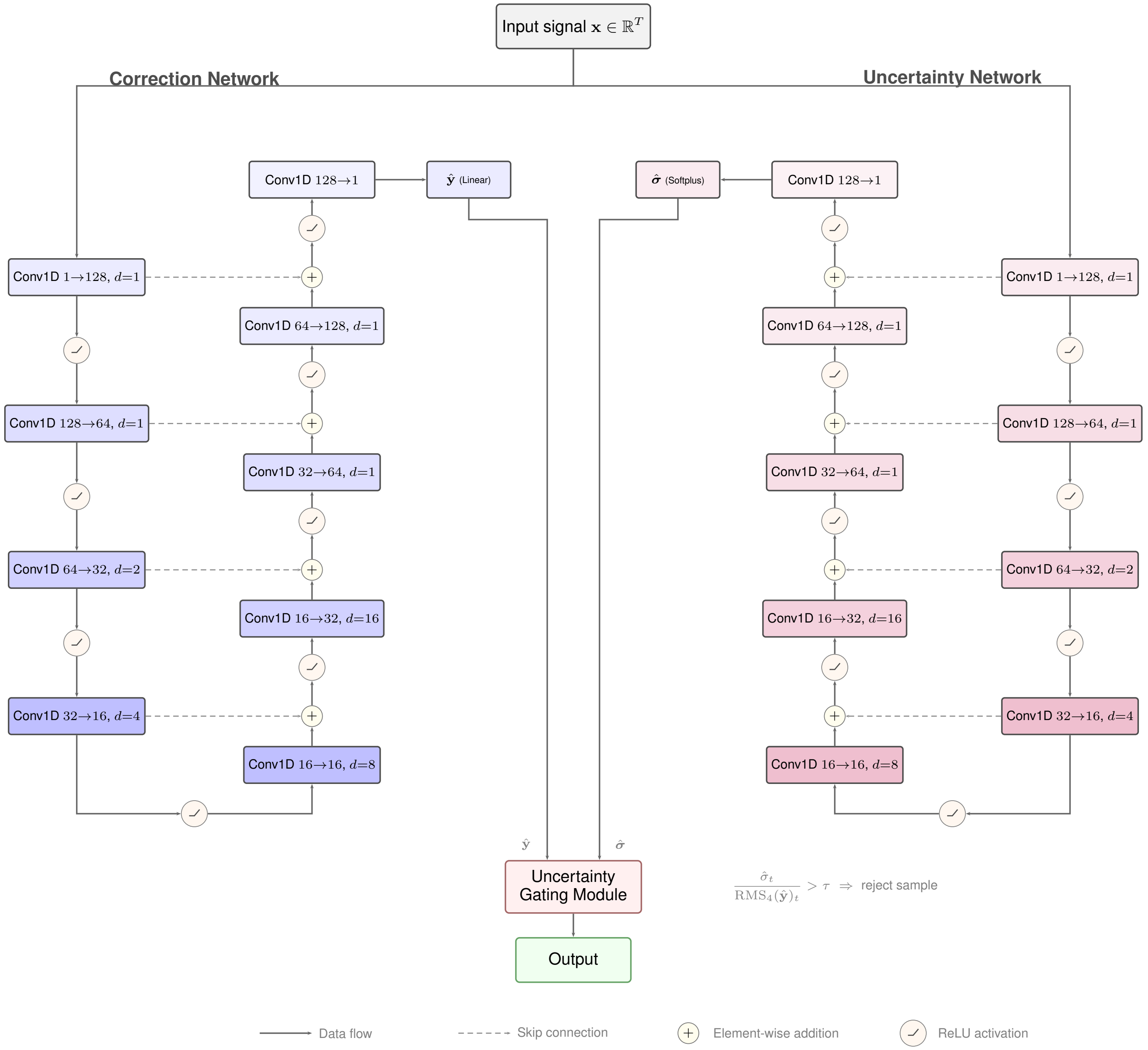


Fig. 1. **Dual-branch neural network architecture.** The system comprises two parallel 1D convolutional networks with identical U-shaped architectures (9 layers each, kernel size $k$=25, same padding) but distinct output activations. Both networks independently process single-channel BSGM input ($\mathbf{x} \in \mathbb{R}^T$). The downstream path uses increasing dilation rates ($d$), expanding the receptive field to capture both short-duration artifact spikes and long-period gastric rhythms. In the upstream path, each convolutional layer's output is summed element-wise with the corresponding encoder layer's pre-activation output (dashed skip connections), preserving fine-grained temporal features. The correction network outputs an estimated clean signal $\hat{\mathbf{y}}$ via a linear activation, while the uncertainty network outputs pointwise aleatoric uncertainty $\hat{\boldsymbol{\sigma}}$ via a Softplus activation. An uncertainty gating module compares each sample's predicted standard deviation to the local signal power, computed as the root-mean-square of the corrected signal over a sliding window of width $w$ ($w$ = 240 s). Samples for which $\hat{\sigma}_t/\mathrm{RMS}_w(\hat{\mathbf{y}})_t > \tau$ ($\tau = 1.25$) are rejected from downstream analysis.

### III. Gastric Emptying Breath Test Recordings

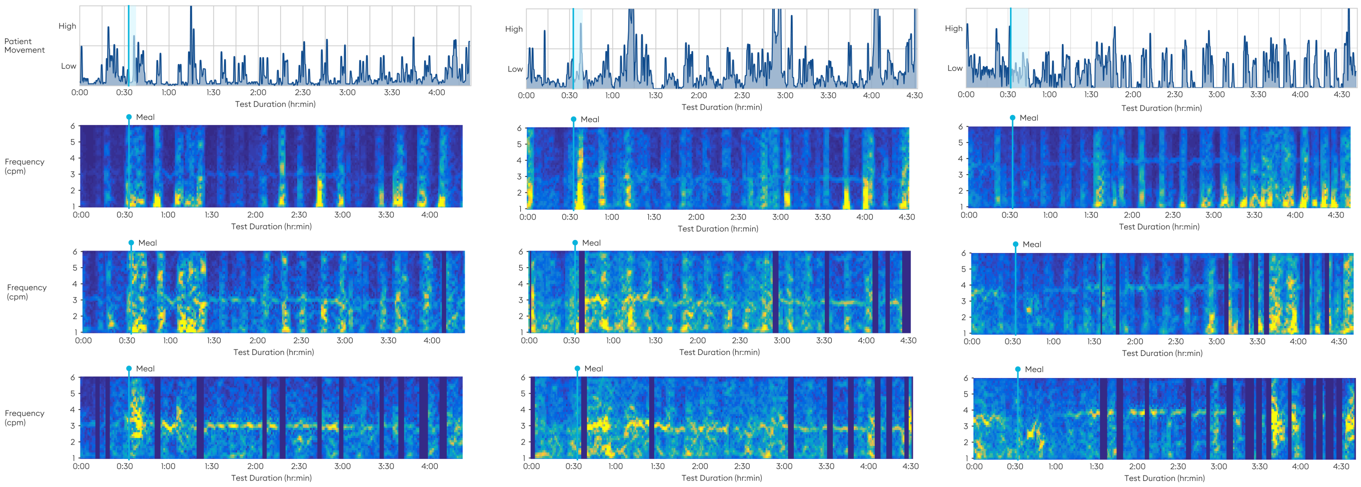


Fig. 2. **Spectrograms and accelerometer-based activity index.** Comparison between artifact filtering methods for three tests (columns) in a research study where the patient was standing up every 15 minutes to perform a gastric emptying breath test. For each test, spectrograms are generated using the complete Gastric Alimetry Algorithm v3.0.0 signal processing pipeline executed with no artifact correction (top), the WF (middle), and the NNF (bottom).

#### IV. 24-Hour Ambulatory Recording

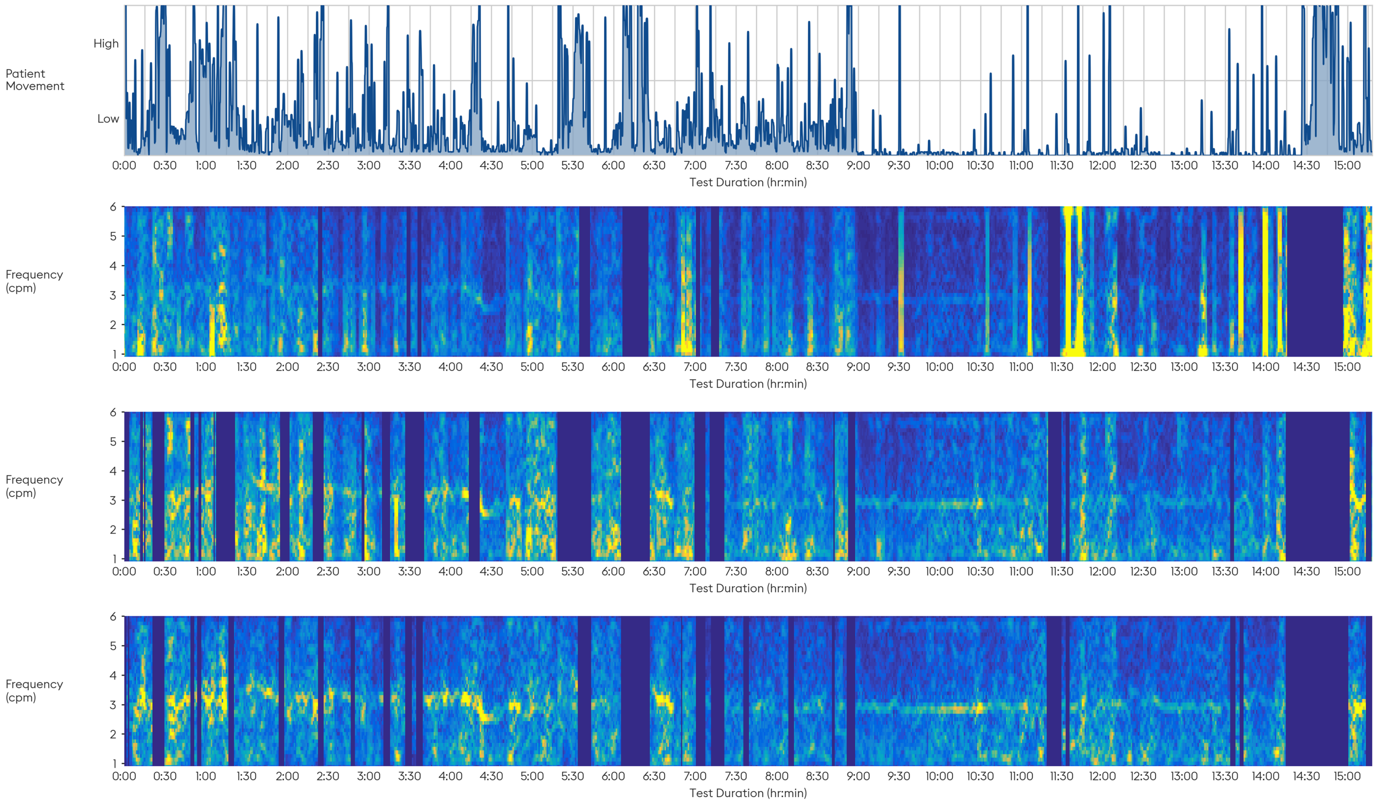


Fig. 3. **Spectrograms and accelerometer-based activity index for a 24-hour recording.** The patient was not limited in their movement throughout the day. Spectrograms are generated using the complete Gastric Alimetry Algorithm v3.0.0 signal processing pipeline executed with no artifact correction (top), the WF (middle), and the NNF (bottom). Aside from the artifact correction module, there are no other modifications to the signal processing between the methods.

#### V. Training and Tuning Cohort Demographics

TABLE I
Demographics, clinical scores, and recording characteristics of the training and tuning cohorts. Continuous variables: Mean ± SD [Range]; categorical: n (%).

| Characteristic | Training Cohort ($N = 2{,}158$) | Tuning Cohort ($N = 240$) |
|---|---|---|
| **Patient Demographics** | | |
| Age (years) | 36.66 ± 16.68 [5.00 – 88.00] | 38.24 ± 17.24 [10.00 – 87.00] |
| Female sex, n (%) | 1,474 (68%) | 174 (72%) |
| BMI (kg/m$^2$) | 25.17 ± 6.20 [12.70 – 70.70] | 24.45 ± 5.67 [12.90 – 42.90] |
| **Patient Symptoms** | | |
| Total Symptom Burden Score | 10.92 ± 13.05 [0.00 – 69.80] | 12.43 ± 13.99 [0.00 – 57.40] |
| Total AGBW Score | 16.61 ± 8.18 [3.00 – 34.00] | 15.50 ± 8.91 [0.00 – 40.00] |
| **Geographical Distribution** | | |
| New Zealand, n (%) | 1,324 (61%) | 93 (39%) |
| Australia, n (%) | 309 (14%) | 22 (9%) |
| Belgium, n (%) | 233 (11%) | 26 (11%) |
| United States, n (%) | 179 (8%) | 74 (31%) |
| Canada, n (%) | 84 (4%) | 9 (4%) |
| United Kingdom, n (%) | 29 (1%) | 9 (4%) |
| Switzerland, n (%) | — | 7 (3%) |
| **Baseline Recording Characteristics** | | |
| Percent Artifact (%) | 26.23 ± 18.96 [0.00 – 100.00] | 24.81 ± 18.80 [0.00 – 97.80] |
| Average Impedance (kΩ) | 134.80 ± 106.83 [11.00 – 849.30] | 136.75 ± 102.34 [21.40 – 638.60] |
| Test Duration (minutes) | 209.00 ± 73.39 [4.00 – 316.00] | 208.21 ± 63.78 [11.00 – 246.00] |

*Repeat testing.* The split between training, tuning and test data is by test and by date (training to 20 November 2023; tuning 22 November 2023 to 6 April 2024; testing after 6 April 2024). Patient identifiers are removed during deidentification, so patient-level separation cannot be verified directly. To estimate how often patients are re-tested, consented commercial tests with age, sex and BMI recorded ($n = 848$ tests at 19 sites with at least two tests) were paired within site where sex matched, BMI agreed within 1.5 kg/m$^2$, height within 3 cm, the recordings did not overlap, and the age difference was consistent with the gap of years between tests. Linking these pairs placed an upper bound of 72 of 848 tests (8.5%) on same-site repeat testing. The number of pairs expected by chance was estimated from the site-level distribution of age differences among candidate pairs that matched on the other criteria (10 rounds of iteration; standard error by delete-a-block jackknife over 10 blocks; the procedure was checked on synthetic data). This gave 11.3 repeat pairs in excess of chance (SE 13.1), or 1.3% of tests (95% CI 0–4.4%). The excess was confined to tests less than 30 days apart (same day, 2.9 pairs; 1–30 days, 6.7); at longer gaps matches occurred at the rate expected by chance (30–180 days, $-0.1$; 180–365 days, 1.4; over one year, 0.5). Because training data end at least 138 days before any test recording, repeat tests of the kind observed are not expected to cross the training–test boundary. The estimate covers consented commercial tests and same-site repeats only; research recordings, which make up most of the training set, and cross-site repeats are not covered. The tuning–test boundary has no gap, but the tuning set was used for epoch selection and threshold review, not for fitting weights. In addition, 22 of the 127 test recordings came from five sites that contributed no training or tuning data.

#### VI. Phenotypic Reclassification Details

Table II lists all 20 tests whose dominant ACv1 phenotypes changed between filters, with the biomarkers under each. Tests 1–9 are the robust reclassifications and tests 10–20 the borderline ones (definitions in Section II-J of the main text); within each group tests are ordered by WF GA-RI. Figs. 4 and 5 show tests 1–9.

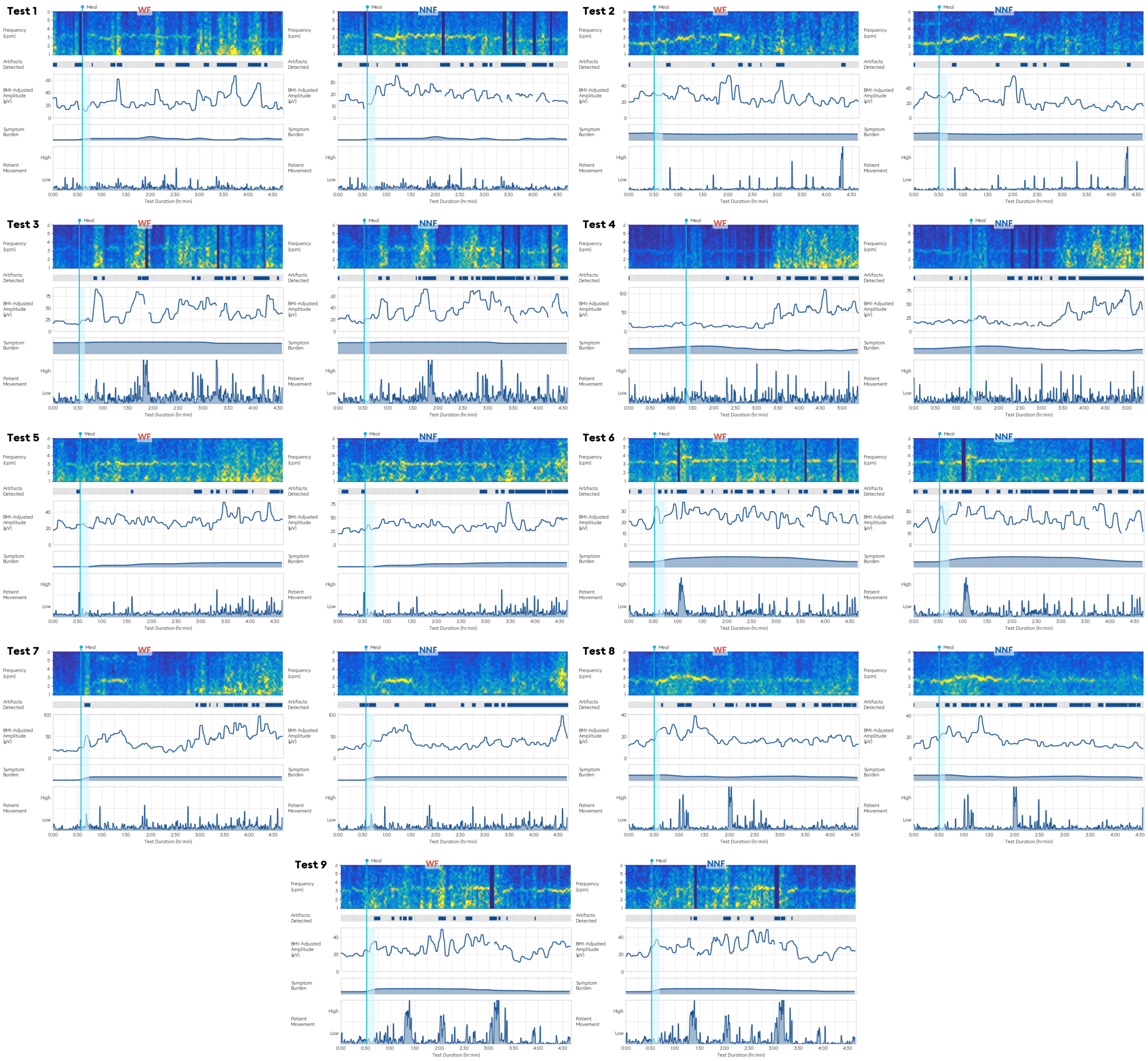


Fig. 4. **Spectrogram comparisons for the 9 tests with robust phenotypic reclassification.** Each panel shows the spectrogram, detected artifacts, BMI-adjusted amplitude, symptom burden, and patient movement traces for the WF-processed (left) and NNF-processed (right) data. Tests are ordered by WF GA-RI value (ascending). Broadband vertical artifacts in the WF spectrograms correspond to periods of elevated patient movement and are substantially attenuated or corrected in the NNF spectrograms, revealing underlying gastric slow wave activity. In 8 of 9 tests, the WF phenotype included a Dysrhythmic classification that was removed or deprioritized by the NNF.

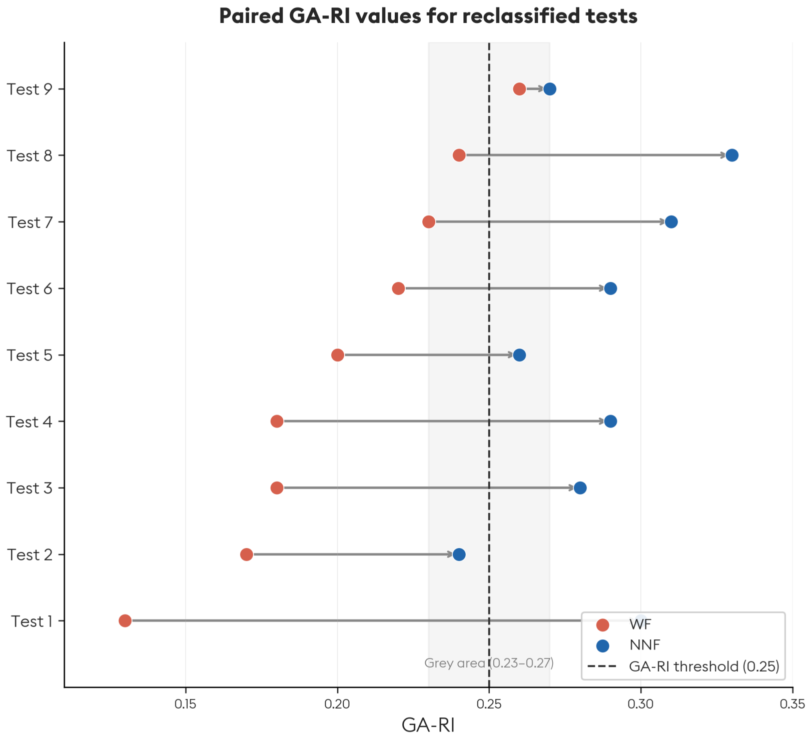


Fig. 5. **Paired GA-RI values for the 9 tests with robust phenotypic reclassification.** Each horizontal bar represents one test, with the WF-derived GA-RI (red) and NNF-derived GA-RI (blue) connected by a directional arrow. The dashed vertical line indicates the normative GA-RI threshold of 0.25, with the shaded region denoting the defined gray area (±0.02). In 7 of 9 tests, the GA-RI shifted from below to above the normative threshold under NNF processing. Tests are ordered by WF GA-RI value (ascending).

TABLE II

ALL 20 TESTS WITH A CHANGE IN DOMINANT ACV1 PHENOTYPES (WF → NNF). GA-RI: GASTRIC ALIMETRY RHYTHM INDEX (NORMATIVE $\geq 0.25$); PGF: PRINCIPAL GASTRIC FREQUENCY (–: UNDEFINED); AMP.: BMI-ADJUSTED AMPLITUDE; MRR: MEAL RESPONSE RATIO (NORMATIVE $\geq 1.0$); ART. DET.: ARTIFACT DETECTED (%)‡.

| Test | ACv1 phenotype(s) | GA-RI | PGF (cpm) | Amp. ($\mu$V) | MRR | Art. det. (%) | Group |
|---|---|---|---|---|---|---|---|
| 1 | Dysrhythmic → Sensorimotor | 0.13 → 0.30 | – → 3.02 | 26.0 → 19.9 | 0.81 → 1.33 | 37.5 → 39.9 | Robust |
| 2 | Dysrhythmic → Dysrh., Continuous | 0.17 → 0.24 | 3.00 → 3.01 | 25.8 → 23.3 | 1.29 → 1.66 | 12.5 → 14.7 | Robust |
| 3 | Dysrhythmic → Continuous | 0.18 → 0.28 | – → – | 40.6 → 36.6 | 0.90 → 0.81 | 25.4 → 48.2 | Robust |
| 4 | Dysrhythmic → Low Meal Response | 0.18 → 0.29 | – → – | 30.7 → 26.8 | 0.25 → 0.36 | 27.2 → 47.8 | Robust |
| 5 | Dysrhythmic → Del. Onset Symptoms | 0.20 → 0.26 | – → 3.00 | 29.2 → 35.6 | 0.86 → 0.91 | 20.4 → 35.4 | Robust |
| 6 | Dysrhythmic → Normal | 0.22 → 0.29 | 3.35 → 3.34 | 25.2 → 23.7 | 1.16 → 1.11 | 35.6 → 54.4 | Robust |
| 7 | Dysrh., Low MR → Normal | 0.23 → 0.31 | – → 2.62 | 41.5 → 37.3 | 0.59 → 1.00 | 26.2 → 48.2 | Robust |
| 8 | Dysrhythmic → Low Meal Response | 0.24 → 0.33 | 2.75 → 2.74 | 18.2 → 16.0 | 1.29 → 1.63 | 40.5 → 56.6 | Robust |
| 9 | Normal → Sensorimotor† | 0.26 → 0.27 | 3.12 → 3.16 | 26.6 → 27.4 | 1.08 → 1.18 | 18.9 → 11.8 | Robust |
| 10 | Dysrhythmic → Dysrh., Sensorimotor | 0.21 → 0.24 | 2.99 → 2.97 | 20.2 → 19.8 | 1.12 → 1.17 | 16.4 → 27.1 | Borderline |
| 11 | Dysrhythmic → Dysrh., Low MR | 0.21 → 0.24 | – → 3.19 | 25.6 → 24.6 | 0.86 → 0.84 | 15.4 → 25.4 | Borderline |
| 12 | Dysrh., Low MR → Dysrhythmic | 0.22 → 0.21 | – → – | 26.5 → 25.5 | 0.88 → 0.91 | 18.8 → 11.2 | Borderline |
| 13 | Dysrh., Sensorimotor → Dysrhythmic | 0.22 → 0.20 | 1.97 → 1.99 | 16.6 → 15.3 | 1.63 → 1.73 | 12.7 → 15.3 | Borderline |
| 14 | Dysrh., Low MR → Low Meal Response | 0.23 → 0.25 | – → 2.95 | 25.7 → 25.2 | 0.92 → 0.95 | 18.2 → 24.5 | Borderline |
| 15 | Sensorimotor, Continuous → Continuous | 0.26 → 0.29 | 2.71 → 2.75 | 25.8 → 24.3 | 0.81 → 0.79 | 34.6 → 42.5 | Borderline |
| 16 | Sensorimotor, Continuous → Continuous | 0.29 → 0.28 | 2.96 → 2.97 | 27.3 → 28.5 | 0.93 → 0.93 | 7.9 → 6.4 | Borderline |
| 17 | Sensorimotor → Low Meal Response | 0.37 → 0.44 | 2.92 → 2.93 | 23.2 → 22.3 | 1.38 → 1.28 | 51.4 → 56.5 | Borderline |
| 18 | Sensorimotor, Del. Onset → Del. Onset Symptoms | 0.43 → 0.42 | 3.14 → 3.12 | 27.2 → 29.7 | 1.14 → 1.04 | 16.2 → 6.8 | Borderline |
| 19 | Low Meal Response → Normal | 0.46 → 0.43 | 3.23 → 3.23 | 32.1 → 35.5 | 0.94 → 1.00 | 7.2 → 7.2 | Borderline |
| 20 | Del. Onset → Sensorimotor, Del. Onset | 0.61 → 0.64 | 3.01 → 3.00 | 47.9 → 47.9 | 1.19 → 1.10 | 26.3 → 34.9 | Borderline |

Tests 1–9, mean ± SD: GA-RI 0.201 ± 0.040 → 0.286 ± 0.027; MRR 0.91 ± 0.34 → 1.11 ± 0.41; Art. det. 27.1 ± 9.3 → 39.7 ± 16.3%.

† Reclassification driven by heartburn correlation crossing the 0.5 threshold (WF: 0.31, NNF: 0.52).

‡ The algorithm's artifact-detected percentage: time points at which the processed signal differs substantially from the raw signal. It rises when more of the signal is corrected and is not the percentage of data removed.

Phenotypes are listed as reported by ACv1. Dysrh.: Dysrhythmic; Low MR: Low Meal Response; Del. Onset: Delayed Onset Symptoms; Normal: no phenotype assigned.

#### VII. Behavior of the Scaled-L1 Loss

Fig. 6 shows the scaling factor $S_t$ (Eq. 2 of the main text) and the resulting per-sample loss for the values used to train the deployed correction network ($N_{\max} = 6$, $R_{\max} = 0.9$). Both were set by design judgment and were not tuned. $N_{\max} = 2$ and $4$ are drawn for illustration only and were not used in training. The weight halves at about 60 times the signal RMS and reaches its floor at about 110 times. Fig. 7 shows the weighting applied to real test samples. For most artifact time points the weight stays close to 1, and it is mainly very large artifacts that are down-weighted. The scaling therefore prevents extreme, unrecoverable artifacts from dominating the gradient while leaving the loss essentially unscaled for the artifacts the network is expected to correct. The sensitivity of the trained network to $N_{\max}$ and $R_{\max}$ was not studied.

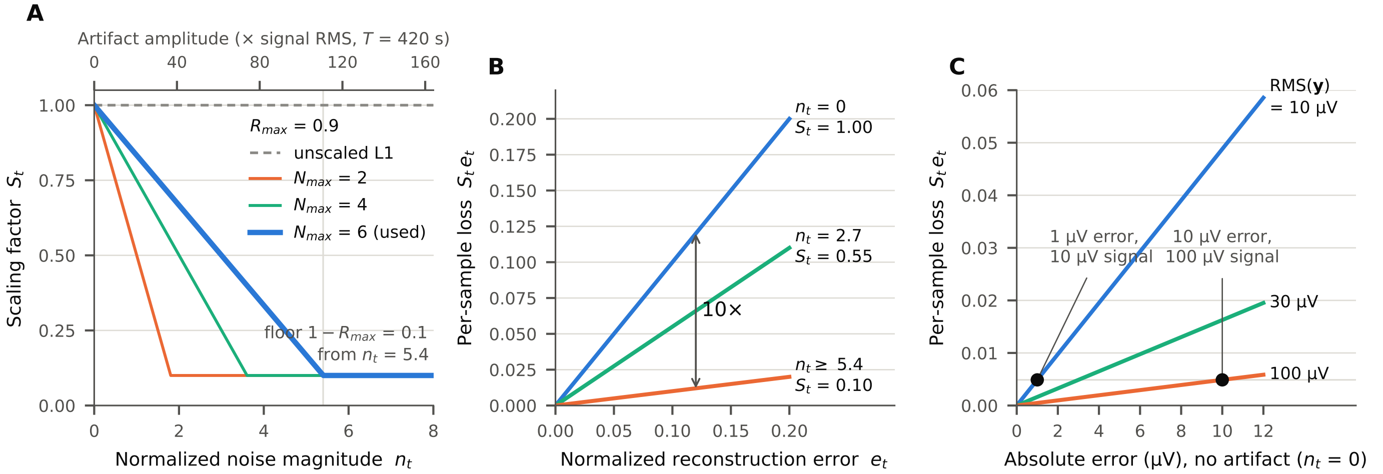


Fig. 6. **Behavior of the amplitude-normalized scaled-L1 loss.** All quantities are normalized by the L2 norm of the clean segment, $\|\mathbf{y}\|_2 = \sqrt{T}\,\mathrm{RMS}(\mathbf{y})$. For the 7-minute training segments at 1 Hz, $T = 420$ and $\|\mathbf{y}\|_2 \approx 20.5\,\mathrm{RMS}(\mathbf{y})$. **(A)** Scaling factor $S_t$ versus normalized noise magnitude $n_t = |x_t - y_t|/\|\mathbf{y}\|_2$ for the values used in this study ($N_{\max} = 6$, $R_{\max} = 0.9$; thick line), with $N_{\max} = 2$ and 4 shown for comparison. $S_t$ falls linearly from 1 at artifact-free time points and reaches its floor of $1 - R_{\max} = 0.1$ at $n_t = N_{\max} R_{\max} = 5.4$, an instantaneous artifact amplitude of roughly 110 times the signal RMS (upper axis). **(B)** Per-sample contribution to $L_c$ (Eq. 1), $S_t\, e_t$ with $e_t = |\hat{y}_t - y_t|/\|\mathbf{y}\|_2$, versus reconstruction error at three noise levels. At equal error, an artifact-free time point contributes ten times the loss of a time point at the noise floor. **(C)** Effect of the $\|\mathbf{y}\|_2$ normalization at artifact-free time points: a 1 $\mu$V error on a 10 $\mu$V-RMS signal incurs the same loss as a 10 $\mu$V error on a 100 $\mu$V-RMS signal, so low- and high-amplitude recordings are weighted equally during training.

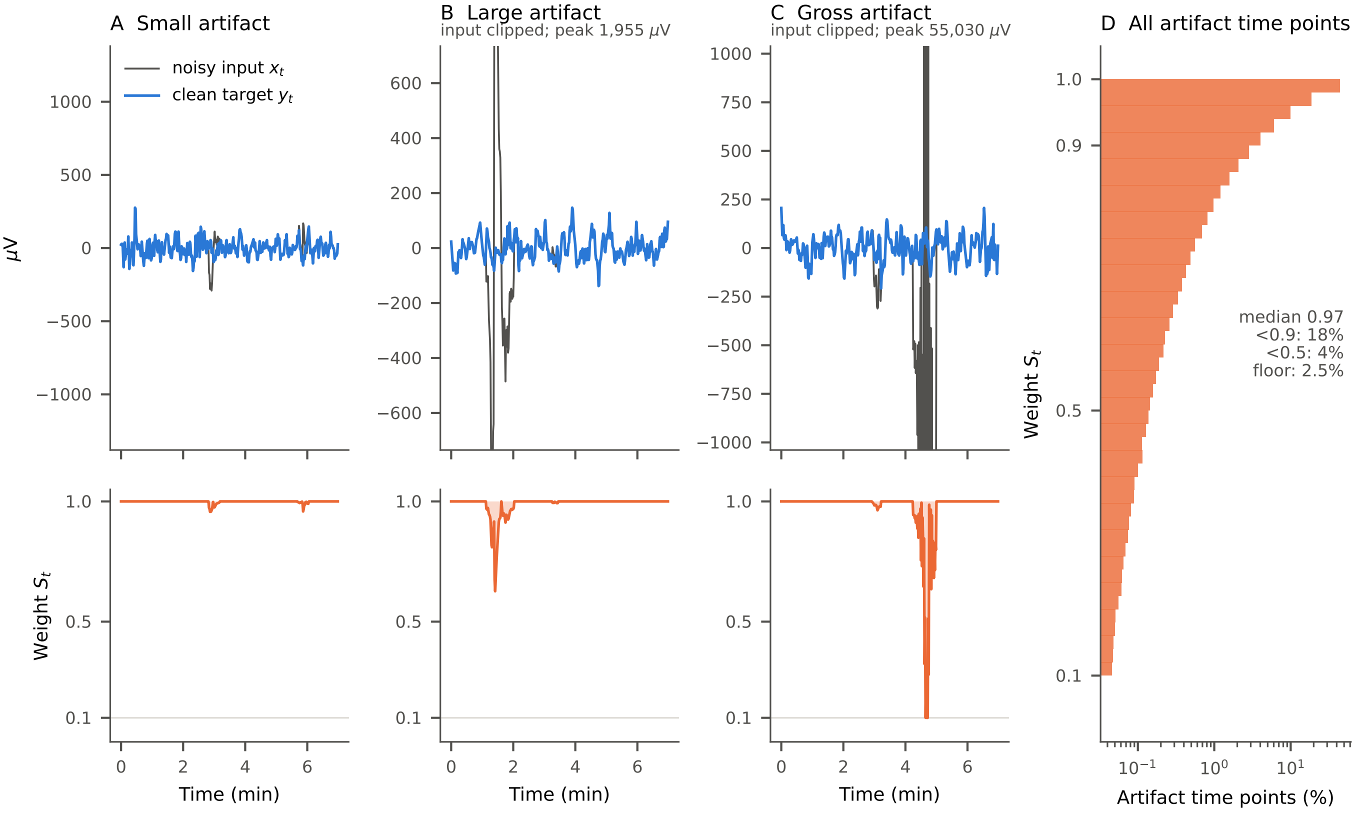


Fig. 7. **The scaled-L1 weighting on real data.** Real test samples, constructed as in training from clean and artifact segments of the same recording (first 7 minutes, the training segment length). **(A–C)** Noisy input $x_t$ and clean target $y_t$ (top; inputs clipped where noted) and the weight $S_t$ applied to the reconstruction error at each time point (bottom) for a small, a large and a gross artifact. **(D)** Distribution of $S_t$ over all 1,419,375 artifact time points in the 37,175 test samples from the 127 test recordings: median 0.97; below 0.9 at 18%, below 0.5 at 4%, and at the floor of 0.1 at 2.5% of artifact time points.

#### VIII. Rejection Stage and Comparators

All analyses in this section use the test samples of main-text Table II (37,175 samples; the two samples on which the WF failed are excluded, as in Table II) and the locked, deployed networks. Nothing was retrained or tuned on test data.

##### A. Gating threshold

Fig. 8 shows segment-level MAPE and data removed as $\tau$ varies from 0.25 to 10 and with gating disabled ($\tau = \infty$). The trade-off is smooth, with no knee that would favor a different value. At the deployed $\tau = 1.25$ the NNF removes 10.2% of data (WF 11.3%) with MAPE 10.0% (WF 16.4%).

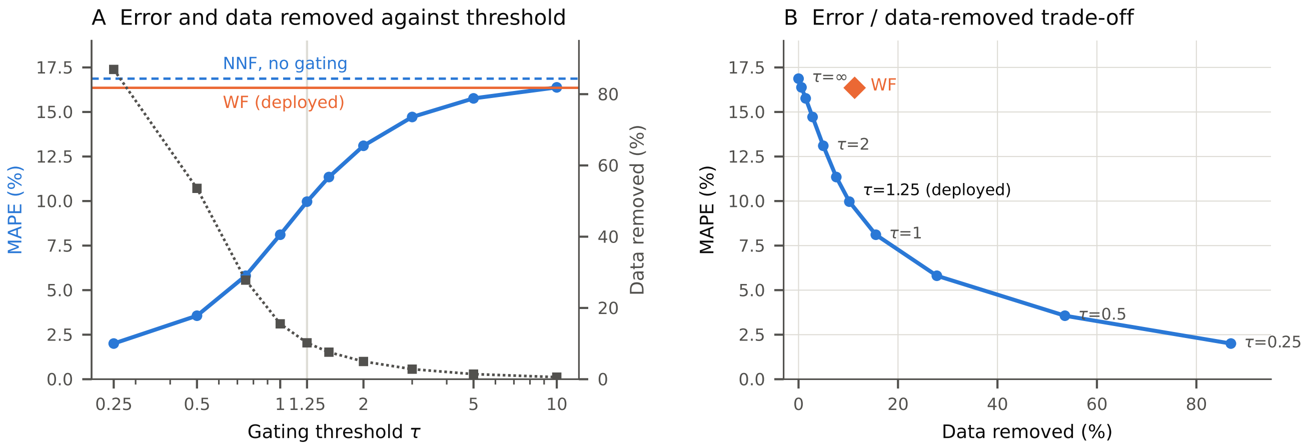


Fig. 8. **Effect of the gating threshold $\tau$ on the test set.** **(A)** Segment-level MAPE (blue, left axis) and percentage of data removed (gray, right axis) against $\tau$; horizontal lines show the NNF with gating disabled (dashed) and the deployed WF (orange). **(B)** The same results as an error–data-removed trade-off, with the WF operating point. MAPE at each $\tau$ (data removed): 0.25, 2.0% (86.9%); 0.5, 3.6% (53.6%); 1, 8.1% (15.5%); 1.25, 10.0% (10.2%); 2, 13.1% (5.0%); 5, 15.8% (1.4%); $\infty$, 16.9% (0%). WF: 16.4% (11.3%).

#### *B. Rejection stage and untrained comparators*

Two untrained single-channel methods were applied to the same samples: a Hampel identifier [3] (121-s rolling median, samples more than 3 robust standard deviations from the median replaced by it) followed by a 1–6 cpm zero-phase bandpass filter (4th-order Butterworth, forward–backward), and wavelet shrinkage (VisuShrink [4], sym8, up to 6 levels, universal threshold with noise level from the finest detail coefficients, hard thresholding). Each was evaluated as a filter alone and with a rejection stage. The rejection score was the 60-s rolling median of $|x_t - f_t|$ between input and filter output, divided by a robust scale of that difference within the sample, and a single global threshold per method was set so that the mean fraction removed equaled that of the NNF (10.22%). Results are given in Table III.

TABLE III
FILTERS WITH AND WITHOUT A REJECTION STAGE ON THE TEST SAMPLES OF MAIN-TEXT TABLE II. MAPE AS MEAN (SD) OVER SAMPLES. PATIENT-LEVEL MAPE AS THE MEAN OF PER-PATIENT MEANS. FILTER-ONLY ROWS ARE COMPARED WITH THE NNF CORRECTION NETWORK WITHOUT GATING, ROWS WITH A REJECTION STAGE WITH THE DEPLOYED NNF (WILCOXON SIGNED-RANK ON PER-PATIENT MEANS, $n = 127$).

| **Method** | **MAPE (%)** | **Removed (%)** | **Patient MAPE (%)** | **NNF better (of 127)** | **p** |
|---|---|---|---|---|---|
| *No rejection stage* | | | | | |
| Unfiltered input | 426.4 (4352.2) | 0 | 350.1 | 127 | $1.4 \times 10^{-22}$ |
| Hampel + bandpass | 202.2 (1933.1) | 0 | 177.7 | 127 | $1.4 \times 10^{-22}$ |
| Wavelet shrinkage | 458.8 (4321.0) | 0 | 383.0 | 127 | $1.4 \times 10^{-22}$ |
| WF, no removal | 28.6 (32.2) | 0 | 27.1 | 121 | $4.6 \times 10^{-22}$ |
| NNF correction network, no gating | 16.9 (26.3) | 0 | 16.9 | — | — |
| *With a rejection stage* | | | | | |
| Hampel + bandpass, matched removal | 82.2 (1155.8) | 10.2 | 76.6 | 127 | $1.4 \times 10^{-22}$ |
| Wavelet shrinkage, matched removal | 625.4 (6102.0) | 10.2 | 518.2 | 127 | $1.4 \times 10^{-22}$ |
| WF (deployed, own artifact mask) | 16.4 (38.5) | 11.3 | 14.6 | 103 | $1.1 \times 10^{-11}$ |
| NNF correction network, WF artifact mask | 9.7 (12.1) | 11.3 | 9.8 | — | — |
| NNF (deployed, $\tau = 1.25$) | 10.0 (30.8) | 10.2 | 10.3 | — | — |

#### IX. Data Preparation Details

*Segment counts.* After preprocessing to separate data into single-channel segments classified as clean (7-minute segments) or noisy (arbitrary length), the training cohort yielded 68,993 clean segments and 4,806,804 noisy segments (reflecting the short, variable duration of individual noise events), and the tuning cohort 9,420 clean and 570,672 noisy segments. Clean and noisy segments were recombined and supplemented with synthetic data to generate training and tuning samples. Noise segments were required to meet a minimum duration and peak absolute amplitude for use in training/tuning.

*Role of the WF.* The foremost priority is ensuring that segments labeled as clean are of high fidelity. Because these segments serve as the ground truth signal for the network, a conservative approach is essential to ensure they truly represent gastric rhythms without spurious artifacts. This method does not assume the WF is a perfect corrector; if it were, the new NNF would be redundant. Instead, the WF is employed simply to extract a plausible noise signature for the network to learn from. While clean gastric signals have specific (though variable) frequency and amplitude profiles, real-world artifacts typically exhibit wide spectral and amplitude variation. Therefore, an imperfect noise estimation by the WF is sufficient to capture the realistic features of noise that the new network is designed to correct.

*Synthetic signals.* To ensure generalization, the real-world data were supplemented with a comprehensive library of synthetically generated data. Expanding upon recent literature demonstrating the value of data augmentation for generating synthetic electrogastrogram time series [5], we developed a stochastic modeling approach to generate synthetic waveforms that capture the physiological variability of gastric myoelectric activity. Synthetic gastric slow waves were modeled as sinusoidal oscillators with non-stationary parameters sampled from empirically-derived probability distributions. To simulate natural physiological variability, we introduced continuous phase drifts (simulating frequency wobble) and amplitude modulation via cumulative random walk processes. While baseline frequencies were sampled from the physiological gastric range (1.5–5.0 cpm), we also generated a subset of "high-scatter" signals by superimposing multiple oscillators with randomized frequencies and phases.

*Synthetic artifacts.* Synthetic artifacts were designed to mimic the challenging temporal and spectral characteristics of patient motion and electrode contact noise, modeled as transient bursts with variable durations (5 seconds to 5 minutes) and randomized onset/offset envelopes. The artifacts were scaled to realistic noise-to-signal ratios, with peak amplitudes reaching orders of magnitude higher than the underlying physiological signal. A subset of synthetic noise segments underwent low-pass filtering with a random 1–6 cpm cutoff to simulate "hard" artifacts that spectrally overlap with the gastric frequency band, forcing the network to learn morphological rather than just spectral discrimination.

*Combining real and synthetic data.* A comprehensive training dataset was constructed by cross-combining real and synthetic sources equally across four permutations (real/synthetic signal $\times$ real/synthetic noise). This hybrid approach allows the networks to anchor on the ground-truth complexity of real patient data while benefiting from the controlled diversity and corner-case coverage provided by the synthetic generation pipeline. Recent work has demonstrated that supplementing real datasets with synthetic examples can significantly enhance model resilience to out-of-distribution artifacts, even if the synthetic examples are not perfectly realistic [6]. When augmenting clean segments with noise, a random number of noise segments (chosen uniformly from 0 to 5) was added, with noise segments distributed throughout the clean segment. The variable number of noise segments was to promote the network learning to correct signals with different degrees of corruption, including signals with no noise so as to ensure the network's ability to preserve clean signals.
